# The anti-tumour effect of CD8^+^ infiltration is associated with Human Leukocyte Antigen Class I expression and tumour proliferation in colorectal cancer

**DOI:** 10.1101/2020.03.25.20036970

**Authors:** Arfon G M T Powell, Colin Richards, Jonathan J Platt, Clare Orange, Lindsay Bennett, Donald C McMillan, Paul G Horgan, Joanne Edwards

## Abstract

**Purpose:** The host inflammatory response is an important determinant of cancer outcome, however, the factor/s that regulate this response remains unclear. We aimed to determine if Human Leukocyte Antigen (HLA) class I and tumour cell proliferation are associated with CD8^+^ infiltration and survival in patients undergoing potentially curative resection for colorectal cancer.

**Methods:** HLA class I expression (W6/32 and B2-microglobulin) and tumour proliferation index (Ki67) were quantified using immunohistochemistry on tissue micro arrays (TMA). The local inflammatory response at the invasive margin was assessed using the Klintrup-Makinen (K-M) score and CD8^+^ infiltration was assessed at the invasive margin (mCD8^+^), stroma (sCD8^+^) and cancer cell nests (cCD8^+^).

**Results:** Preserved HLA class I expression was associated with lower Dukes’ stage (p=0.032), lower T stage (p=0.040) and higher cCD8^+^ (p=0.003). High Ki67 was associated with a good K-M score (p<0.001), higher mCD8^+^ (p=0.033), higher sCD8^+^ (p=0.025) and higher cCD8^+^ (p<0.001). On binary logistical regression analysis both preserved HLA class I expression (HR 1.99 95%CI (1.13–3.51),p=0.012) and high Ki67 (HR 2.63 95%CI (1.08–6.38),p=0.033) were independently associated with higher CD8^+^ infiltration within the cancer cell nests. On multivariate survival analysis, preserved HLA class I expression was associated with disease free (HR 0.47 95%CI (0.25–0.89),p=0.020) and cancer specific survival (HR 0.52 95%CI (0.28–0.97),p=0.038).

**Conclusion:** This study suggests that a pronounced local inflammatory response is independently associated with both, HLA class I expression and tumour proliferation.

## Introduction

Colorectal cancer (CRC) is the second most common cause of cancer death in Western Europe and North America. Each year in the UK, there are approximately 35,000 new cases and 16,000 deaths attributable to this disease.[1] Overall survival is poor; even in those who undergo resection with curative intent; approximately a third will develop recurrence and die of their disease.[2] Adjuvant chemotherapy is associated with significant morbidity and results from trials suggest no survival benefit for patients with node negative disease. Subsequently, there has been a considerable effort to develop and validate tests that aid identification of high-risk patients to enable risk stratification.

Disease progression and metastasis is dependent upon a complex relationship between tumour behaviour and the anti-tumour effects of the local inflammatory response.[3] A pronounced local immune response at the invasive margin, which can be measured by cytotoxic CD8^+^ lymphocyte infiltration and the Klintrup–Makinen score (K-M) has been associated with improved stage independent survival. [3,4]

The factors that regulate the local immune response remains unclear, however, tumour antigen presentation via the Human Leukocyte Antigen (HLA) class I complex has been suggested. Loss of HLA class I expression has been associated with reduced CD8^+^ infiltration, tumour progression and metastasis, however, studies reporting the prognostic value of HLA class I are few in number with conflicting reports. [5]

Tumours with mismatch repair (MMR) protein deficiency have a pronounced CD8+ lymphocyte infiltration, high levels of proliferation and have complete loss of HLA class I expression. [6] It remains unclear if HLA class I and proliferation are independently associated with a pronounced inflammatory response. The aim of the present study was to determine if HLA class I and tumour proliferation are independently associated with a pronounced local inflammatory response and survival in patients undergoing potentially curative resection for colon and rectal cancer.

## Material and Methods

### Patients

All patients with histologically proven CRC who, on the basis of pre-operative staging CT scans and findings at laparotomy, were considered to have undergone potentially curative resection for CRC (Stages I–III) between 1997 and 2005 in a single surgical unit were included in the study. Patients who had either pre-operative chemo-radiotherapy or active chronic inflammatory disease were excluded. Patients who died within 30 days of surgery were excluded from the survival analysis. The tumours were staged according to conventional TNM classification and additional pathological characteristics were recorded from pathology reports issued at the time of resection. Tumours proximal to the rectosigmoid junction were considered colonic and those distal to and including the rectosigmoid junction were considered rectal tumours.[7] Lymph node ratio was already available and described previously. [8]

Adjuvant therapy was given to patients following discussion at the multidisciplinary team meeting. Patients’ clinical and pathological information was available to the oncologist making these decisions and the treatment offered was based on treatment guidelines for CRC at that time. This study was approved by the research ethics committee, Glasgow Royal Infirmary, Glasgow. Consent for tissue use was obtained at the time of consenting for the surgical procedure.

## Methods

### Tissue micro array (TMA) construction

In brief, each specimen had tumour rich area identified by a pathologist and four 0.6-mm^2^ tumour cores were used to construct the TMA.[9] These were used to assess Ki67 and HLA class I (W6/32 and β2-microgobulin) expression by immunohistochemistry.

### Immunohistochemistry (IHC)

TMA sections (4um) were cut and mounted on slides coated with aminopropyltriethoxysilane. TMAs were dewaxed and rehydrated. Antigen retrieval was performed by immersing slides in citrate buffer (pH 6.0) for Ki67, Tris-EDTA buffer (pH 8.0) for CD8 and Tris-EDTA buffer (pH 9.0) for W6/32 and β2-microglobulin (B2-m) and microwaved under pressure for 5 minutes and cooled for 20 minutes. Endogenous peroxidise activity was blocked by incubation in 3% H_2_O_2_ for 10 minutes. After transfer to a humidified chamber, sections were blocked by incubation with casein solution (Vector Laboratories, Peterborough, England) at a 1:1 dilution with Tris-buffered saline (TBS) for 1 hour and then primary antibody overnight at 4^°^ C. Dako anti-Ki67 (Monoclonal mouse anti-human, Ki-67 antigen, Clone MIB-1, CodeM7240, DAKO, Glostrup, Denmark) W6/32 (Monoclonal anti-human, HLA class I heavy chain, CodeAB7855, ABCAM, Cambridge, UK) and B2-m (Monoclonal anti-human, HLA class I light chain, CodeAB, ABCAM, Cambridge, UK) were used with dilutions of 1:50 and CD8^+^ (Monoclonal Mouse Anti-Human CD8, Clone C8/144B, CodeM7103, DAKO; Glostrup, Denmark) at a dilution of 1:100. Antibody antigen binding was detected using the peroxidase-based Envision (Dako, Cambridgeshire, UK) technique using a diaminobenzidine chromogenic substrate system. Appropriate positive controls were included in each run. Negative controls were omission of the primary antibody. Sections were then counterstained using Haematoxylin, dehydrated, and mounted in DPX.

#### Slide Scanning and Scoring

Stained slides were scanned using a Hamamatsu NanoZoomer (UK). Visualization and automated assessment were carried out using the Slidepath Tissue IA system version 1.0 (Ireland) and observer assessment of percentage of positive cells was performed on a computer monitor.

### Image Analysis of Ki67 staining

For automated image analysis, digitised slides were accessed through the Slidepath Image Analysis system and evaluated with the program’s nuclear scoring algorithm through quantifying nuclear staining within individual cores and deriving a counting score for each target area. This method has been described previously by Mohammed et al. [10]

### Assessment of tumour proliferation index (PI)

PI represents a positive percentage expression of Ki67 following counting of positive and negative nuclei in each tumour core. High and low expression of Ki67 was based on previously published thresholds.[10] The mean index score for each protein was used as a representative score for each tumour. An observer blinded to patient outcome independently scored 108 cores and the Interclass Correlation Coefficient (ICCC) was 0.94, which indicates excellent agreement.

### Assessment of CD8^+^ expression

Full sections of representative areas of tumour were assessed for CD8^+^ lymphocyte infiltration by IHC.[11] The degree of infiltration was established by counting the number of immune cells exhibiting staining. CD8^+^ infiltration was categorised into 4 groups, 0 (0-3 immune cells exhibiting staining), 1 (3-25 immune cells), 2 (26-50 immune cells) and 3 (greater than 50 cells). Forty sections were independently scored by an observer blinded to patient outcome; ICCC >0.80.

### Assessment of HLA class I expression

HLA class I heavy chain (W6/32) and light chain (B2-m) expression was established by calculating the percentage positive cells exhibiting membrane staining for each antibody. HLA class I expression thresholds were determined via a method previously used by Watson et al.[12] Strong positive staining of fibroblasts and immune cells indicating success of the technique and served as an internal positive control. The mean index score for each protein was used as a representative score for each tumour. An observer blinded to patient outcome independently scored 108 cores with an ICCC 0.84.

### Analysis

The ICCC for each protein was calculated to confirm consistency between observers and the mean of the two observers’ scores were used for analysis. The ICCC values were consistently above 0.8 (ICCC values ≥0.7 were considered acceptable). Inter-relationships between the various clinical parameters were calculated using Pearson’s Chi square test. Because of the number of statistical comparisons, a *P*-value of <0.05 was considered to be significant.

Disease specific survival rates were generated using the Kaplan-Meier method. The log rank test was used to compare significant differences between subgroups using univariate analysis. Multivariate stepwise Cox-regression analysis was performed to identify factors that were independently associated with disease specific death. A stepwise backward procedure was used to derive a final model of the variables that had a significant independent relationship with survival. To remove a variable from the model the corresponding p-value had to be greater than 0.05. The statistical analyses were performed using a statistical software package (SPSS 18.0 Inc., Chicago, IL, USA).

## Results

### IHC analysis of proliferation, apoptosis and survival

#### Clinicopathological Details

The majority of patients were under 75 years of age (70%), male (54%), were node negative (57%), had colon cancer (68%) and 29% received post-operative adjuvant chemotherapy. The minimum follow-up was 46 months; the mean follow-up of the survivors was 112 months. During this period, 70 patients had a cancer related death and 69 patients had a non colorectal cancer death.

### Tumour cellular protein expression

#### Tumour proliferation index (Ki67)

Ninety six percent of patients exhibited nuclear expression. Tumours were subdivided into those with high (>15% positive proliferation) or low (≤15% positive proliferation) expression.

On univariate analysis high Ki67 expression was significantly associated with better disease free (p=0.011, Table 1) and cancer specific survival (p=0.010, Table 2), however this was not independent on multivariate analysis. Chi squared analysis demonstrated that high Ki67 was associated with poor differentiation (p=0.036) and good K-M score (p<0.001) (Table 3).

**Table 1.** **The relationship between clinicopathological characteristics and disease free survival in patients undergoing surgery for colorectal cancer (n= 271): Univariate and multivariate analysis**.

|  |  |  | Univariate analysis |  | Multivariate analysis |
| --- | --- | --- | --- | --- | --- |
|  | Patients<br>(n= 271) | p-value | Hazard ratio<br>(95% CI) | p-value | Hazard ratio<br>(95% CI) |
| Age (<75 / ≥75 years) | 189 / 82 | 0.723 | 1.10 (0.65 – 1.86) |  |  |
| Sex (Female / Male) | 124 / 147 | 0.159 | 1.43 (0.87 - 2.37) |  |  |
| Site (Colon / Rectum) | 184 / 87 | 0.444 | 0.89 (0.66 – 1.20) |  |  |
| T stage (1 / 2 / 3 / 4) | 9 / 21 / 161 / 80 | 0.010 | 1.66 (1.13 – 2.45) | 0.152 |  |
| Lymph node ratio (0 / 0.01–0.24 / 0.25–0.49 / >0.49) | 155 / 86 / 25 / 5 | 0.011 | 1.46 (1.09 – 1.95) | 0.825 |  |
| Lymph node sample (≥12 / <12) | 159 / 112 | 0.189 | 1.38 (0.85 – 2.24) |  |  |
| Differentiation (Mod / Poor) | 240 / 31 | 0.927 | 1.04 (0.47 – 2.27) |  |  |
| Adjuvant therapy (No / Yes) | 192 / 79 | 0.035 | 1.70 (1.04 – 2.79) | 0.533 |  |
| K-M Score (Good / Poor) | 89 / 182 | 0.007 | 2.31 (1.26 – 4.24) | 0.302 |  |
| Ki67 (Low / High) n=267 | 107 / 162 | 0.011 | 0.53 (0.33 – 0.87) | 0.112 |  |
| HLA Class I (Low / Moderate / High) | 73 / 56 / 15 / | 0.013 | 0.48 (0.27 – 0.85) | 0.020 | 0.47 (0.25 – 0.89) |
| mCD8<sup>+</sup> Infiltration (Absent / Low / Intermediate / High) | 49 / 101 / 73 / 25 | 0.001 | 0.60 (0.44 – 0.81) | 0.192 |  |
| sCD8<sup>+</sup> Infiltration (Absent / Low / Intermediate / High) | 74 / 117 / 48 / 17 | 0.009 | 0.65 (0.48 – 0.90) | 0.375 |  |
| cCD8<sup>+</sup> Infiltration (Absent / Low / Intermediate / High) | 85 / 100 / 40 / 32 | 0.001 | 0.59 (0.44 – 0.80) | 0.025 | 0.63 (0.42 – 0.94) |

**Table 2.** **The relationship between clinicopathological characteristics and cancer specific survival in patients undergoing surgery for colorectal cancer (n= 271): Univariate and multivariate analysis**.

|  |  |  | Univariate analysis |  | Multivariate analysis |
| --- | --- | --- | --- | --- | --- |
|  | Patients<br>(n= 271) | p-value | Hazard ratio<br>(95% CI) | p-value | Hazard ratio<br>(95% CI) |
| Age (<75 / ≥75 years) | 189 / 82 | 0.016 | 1.81 (1.12 – 2.93) | 0.672 |  |
| Sex (Female / Male) | 124 / 147 | 0.498 | 1.18 (0.73 - 1.90) |  |  |
| Site (Colon / Rectum) | 184 / 87 | 0.270 | 0.85(0.64 – 1.14) |  |  |
| T stage (1 / 2 / 3 / 4) | 9 / 21 / 161 / 80 | 0.002 | 1.81 (1.23 – 2.67) | 0.264 |  |
| Lymph node ratio (0 / 0.01–0.24 / 0.25–0.49 / >0.49) | 155 / 86 / 25 / 5 | 0.003 | 1.53 (1.15 – 2.02) | 0.930 |  |
| Lymph node sample (≥ 12 / <12) | 159 / 112 | 0.240 | 1.33 (0.83 – 2.12) |  |  |
| Differentiation (Mod / Poor) | 240 / 31 | 0.384 | 1.37 (0.68 – 2.75) |  |  |
| Adjuvant therapy (No / Yes) | 192 / 79 | 0.360 | 1.26 (0.77 – 2.08) |  |  |
| K-M Score (Good / Poor) | 89 / 182 | <0.001 | 3.31 (1.69 – 6.47) | 0.204 |  |
| Ki67 (Low / High) n=267 | 107 / 160 | 0.010 | 0.54 (0.33 – 0.86) | 0.325 |  |
| HLA Class I (Low / Moderate / High) | 73 / 56 / 15 / | 0.018 | 0.51 (0.29 – 0.89) | 0.038 | 0.52 (0.28 – 0.97) |
| mCD8<sup>+</sup> Infiltration (Absent / Low / Intermediate / High) | 49 / 101 / 73 / 25 | <0.001 | 0.53 (0.39 – 0.72) | 0.044 | 0.65 (0.43 – 0.99) |
| sCD8<sup>+</sup> Infiltration (Absent / Low / Intermediate / High) | 74 / 117 / 48 / 17 | 0.003 | 0.63 (0.46 – 0.86) | 0.568 |  |
| cCD8<sup>+</sup> Infiltration (Absent / Low / Intermediate / High) | 85 / 100 / 40 / 32 | <0.001 | 0.53 (0.39 – 0.71) | 0.071 |  |

**Table 3.** **The relationship between clinicopathological factors and cell signalling pathways in patients undergoing elective surgery for colorectal cancer**

|  | Sex | Site | Dukes | T stage | LNR | Differentiation | K-M Score | Ki67 | HLA | mCD8<sup>+</sup> | sCD8<sup>+</sup> | cCD8<sup>+</sup> |
| --- | --- | --- | --- | --- | --- | --- | --- | --- | --- | --- | --- | --- |
| **Age**<br>(<75 / ≥75) | 0.080 | **0.022\*** | 0.699 | 0.953 | 0.382 | 0.212 | 0.374 | 0.972 | 0.808 | 0.327 | 0.242 | 0.983 |
| **Sex**<br>(Female / Male) |  | 0.275 | 0.917 | 0.894 | 0.518 | 0.944 | 0.391 | 0.498 | 0.015* | 0.282 | 0.630 | 0.972 |
| **Site**<br>(Colon / Rectum) |  |  | 0.753 | **<0.001\*** | 0.656 | 0.426 | 0.643 | 0.628 | 0.199 | 0.750 | 0.693 | 0.662 |
| **Dukes**<br>(A / B / C) |  |  |  | **<0.001** | **<0.001** | 0.059 | 0.057 | 0.620 | **0.012\*** | 0.797 | 0.346 | **0.047\*** |
| **T stage**<br>(1 / 2 / 3 / 4) |  |  |  |  | **0.003** | 0.084 | **0.001** | 0.281 | **0.036\*** | 0.211 | 0.982 | 0.253 |
| **LNR**<br>(0 / 0.01-0.24 / 0.25-0.49 / >0.5) |  |  |  |  |  | 0.138 | 0.478 | 0.722 | 0.061 | 0.887 | 0.414 | **0.036\*** |
| **Differentiation**<br>(Mod / Poor) |  |  |  |  |  |  | 0.266 | **0.036** | 0.307 | 0.714 | 0.254 | 0.039 |
| **K-M Score**<br>(Good / Poor) |  |  |  |  |  |  |  | **<0.001\*** | 0.127 | **<0.001** | **<0.001** | **<0.001** |
| **Ki67**<br>(Low / High) |  |  |  |  |  |  |  |  | 0.240 | **0.033** | **0.025** | **<0.001** |
| **HLA Class I**<br>(Absent/Low/Intermediate/High) |  |  |  |  |  |  |  |  |  | 0.354 | 0.052 | **0.003** |
| **mCD8<sup>+</sup>**<br>(Absent/Low/Intermediate/High) |  |  |  |  |  |  |  |  |  |  | **<0.001** | **<0.001** |
| **sCD8<sup>+</sup>**<br>(Absent/Low/Intermediate/High) |  |  |  |  |  |  |  |  |  |  |  | **<0.001** |
| **cCD8<sup>+</sup>**<br>(Absent/Low/Intermediate/High) |  |  |  |  |  |  |  |  |  |  |  |  |

\*Indicates an inverse relationship

#### Human Leukocyte Antigen (HLA) Class I expression

Membrane expression of W6/32 was seen in 93% of patients and cytoplasmic staining in 96%. Tissue from 236 patients was available for analysis with 12% exhibiting strong staining (>50% cells expressing), 35% moderate staining (26-50% cells expressing), 47% had low staining (1-25% cells expressing) and 7% had no staining. Membrane expression of B2-m was seen in 67% of patients and cytoplasmic staining in 70%. Tissue from 223 patients was available for analysis with 7% exhibiting strong staining (>50% cells expressing), 19% moderate staining (26-50% cells expressing), 41% had low staining (1-25% cells expressing) and 33% had no staining.

For effective antigen presentation, tumour cells require both light and heavy chain sub units. Using the method described by Watson et al, we combined the results of both antibodies to reflect a theoretically functioning HLA complex. Patients were classified as; absent if tumours did not express one or both chains, low expression (low W6/32 and positive B2-m), moderate (moderate W6/32 and positive B2-m) and strong (strong W6/32 and positive B2-m).

On log-rank analysis, high HLA class I expression was significantly associated with better disease free (p=0.047, figure 1) and cancer specific survival (p=0.010). Similar to previous findings we observed that absent HLA class I expression confers an improved survival benefit when compared to low expression (figure 1). Given this finding, only tumours expressing both sub units of the HLA complex were taken into the multivariate model. On multivariate survival analysis high HLA expression was independently associated with disease free (HR 0.47 95%CI (0.25–0.89), p=0.020, table 1) and cancer specific survival (HR 0.52 95%CI (0.28–0.97), p=0.038, table 2).

**Figure 1.**
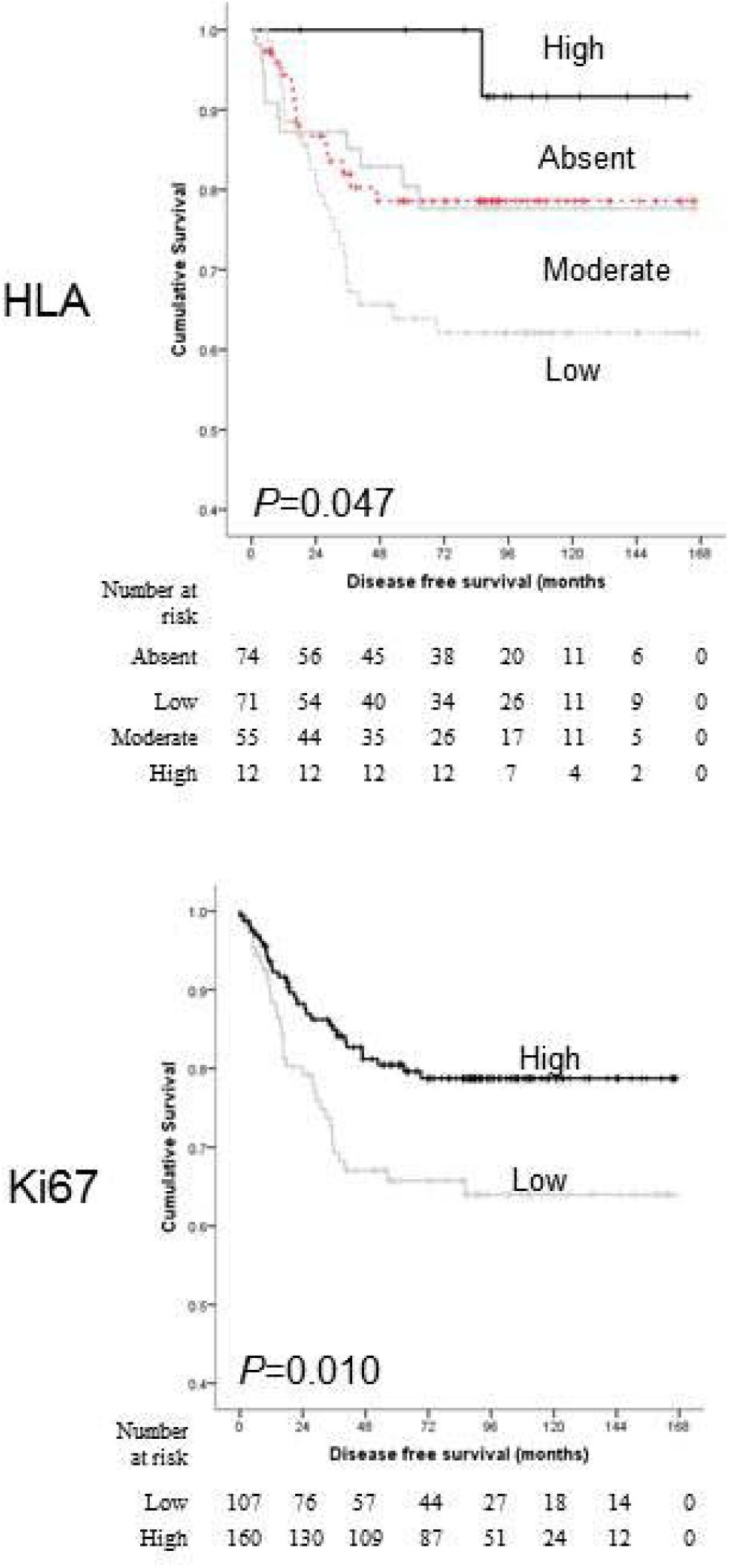
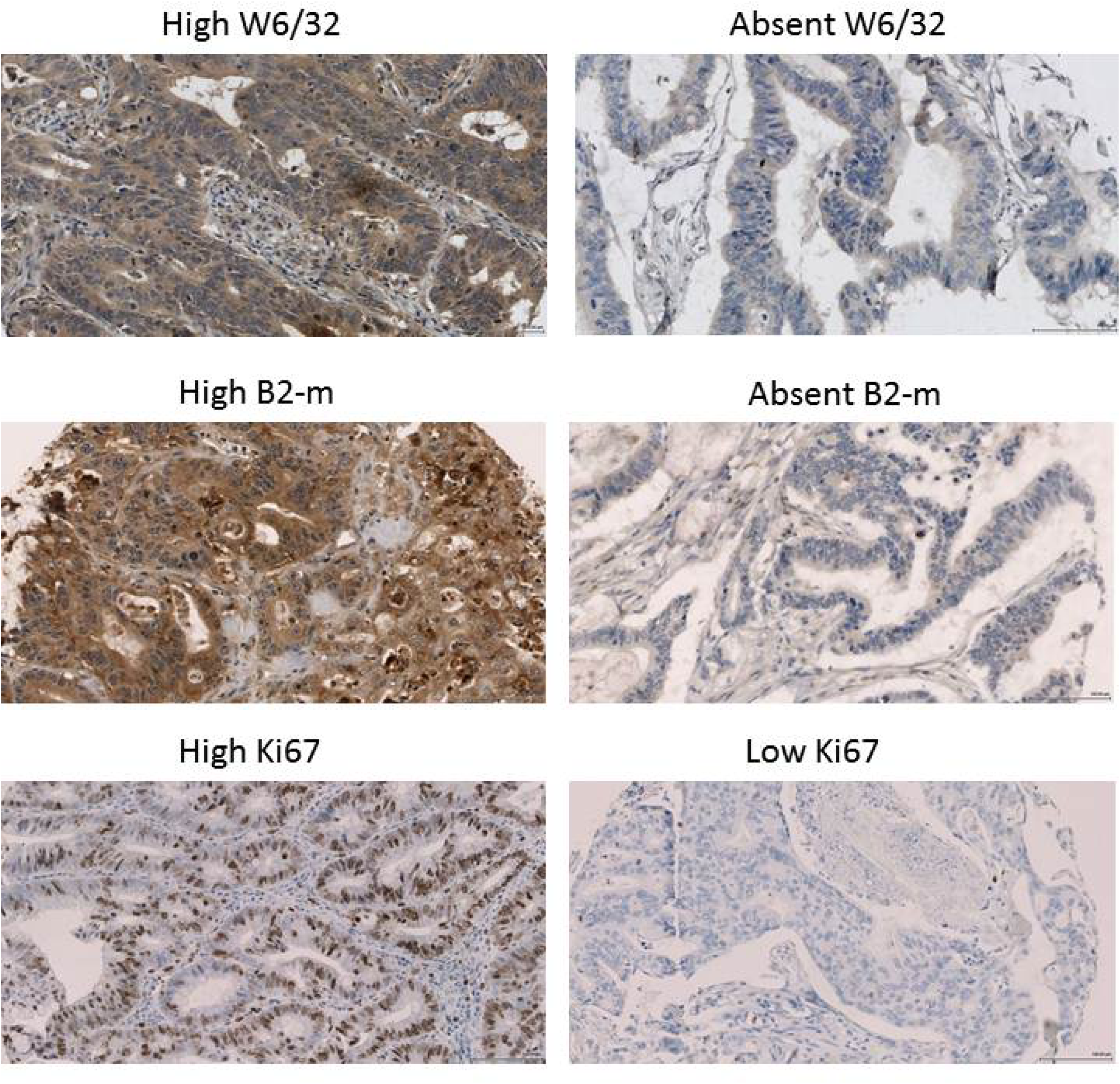
A) The relationship between HLA class I expression, tumour proliferation and disease free survival. B) High and Low staining for W6/32, B2-m and Ki67

Chi squared analysis demonstrated that high HLA class I complex expression correlated with female gender (p=0.015), lower Dukes’ stage (p=0.012), lower T stage (p=0.036), good K-M score (p=0.001) and high cCD8^+^ (p=0.003) (Table 3).

### Tumour CD8^+^ Lymphocyte infiltration

The CD8^+^ lymphocyte infiltrate was measured in three separate areas, the invasive margin (mCD8^+^), intra-tumoral stroma (sCD8^+^) and in the cancer cell nests (cCD8^+^) as described previously (Galon, 2006). Eighty percent of tumours expressed CD8^+^ cells at the invasive margin, 71% expressed CD8^+^ cells in the stroma and 67% expressed CD8^+^ cells in the cancer cell nests. Tumours were divided into absent, low, intermediate and high.

On univariate analysis high mCD8^+^ (p=0.001), high sCD8^+^ (p=0.009) and high cCD8^+^ (p=0.001) were all significantly associated with better disease free and cancer specific survival (p<0.001, p=0.003, p<0.001 respectively; Table 2). On multivariable survival analysis, only high cCD8^+^ (HR 0.63 95%CI (0.42–0.94); p=0.025) independently predicted disease free survival and high mCD8^+^ (HR 0.65 95%CI (0.42–0.99); p=0.044) was independently associated with cancer specific survival.

Chi squared analysis demonstrated that high mCD8^+^ was associated with; good K-M score (p<0.001), high Ki67 (p=0.033), high sCD8^+^ (p<0.001) and high cCD8^+^ (p<0.001). High sCD8^+^ was associated with good K-M score (p<0.001), low CRP (p=0.045), high Ki67 (p=0.025) and high cCD8^+^ (p<0.001). High cCD8^+^ was associated with lower Dukes’ stage (p=0.045), lower LNR (p=0.036), good K-M score (p<0.001), high Ki67 (p<0.001) and high HLA Class I (p=0.003). On binary logistical regression analysis both high HLA class I expression (p=0.012; HR 1.99 95%CI (1.13–3.51)) and high Ki67 (p=0.033; HR 2.63 95%CI (1.08–6.38)) were independently associated with a higher cCD8^+^.

## Discussion

The results of this study further supports previous findings that HLA class I expression, proliferation, and CD8^+^ infiltration are important determinants of survival in patients undergoing potentially curative resection for colorectal cancer. This study suggests that the development of the local inflammatory response may arise via two independent mechanisms, HLA class I expression and tumour proliferation. With the proposed introduction of targeted biological therapies, changing the physiological behaviour of the cancer cell may consequently have far reaching effects on the immune response as well as outcome.

HLA class I expression[4] and proliferation[6] have both been associated with higher levels of tumour infiltrating lymphocytes and improved survival, however, this is the first study to examine both factors simultaneously. The results of this study support previous observations relating to the different mechanisms underpinning the initiation and consolidation of the local immune response. Despite MMR deficient tumours being associated with complete loss of HLA class I expression, they commonly have high levels of Dendritic cells, T-regulatory lymphocytes and CD8^+^ lymphocytes.[13–15] Working within the limitations of these types of studies, there appears to be at least 2 different cancer cell behaviours associated with a pronounced local immune response.

Sustained proliferative signalling and avoiding immune destruction are hallmarks of the colorectal cancer cells’ malignant behaviour. Targeted biological therapies aim to suppress these functions by specifically targeting signalling pathways that control these malignant behaviours.[16] Unfortunately, it is not possible to infer mechanistic information from an association study of this type; however, altering the behaviour of the cancer cell may impact on the quality of the immune response and mechanistic laboratory work is needed.

We observed that the presence of CD8^+^ lymphocytes within the cancer cell nests was an independent predictor of poor survival. This confirms previous observations by Galon and colleagues who observed that the type, density, and location of immune cells in colorectal tumours can provide additional prognostic information.[17] In terms of creating a prognostic model, there is now compelling evidence that the product of inflammation is superior to the factors making up the underlying process. Furthermore, Oginio and colleagues demonstrated that the survival benefit of lymphocyte infiltration was independent of any molecular or genetic features.[18] Unfortunately, they did not specifically study CD8^+^ infiltration and confirmation in an appropriately powered cohort is required to determine its use as a prognostic biomarker. With the inclusion of histopathological features into the 7^th^ edition of the TNM staging system for oesophageal cancer[19], validation of CD8^+^ as a prognostic biomarker may aid its incorporation into future staging revisions for colorectal cancer Therefore, delineating the physiology of the immune response may offer a novel therapeutic target for patients with low CD8^+^ infiltration.

In conclusion, this study demonstrates an independent association between tumour proliferation, HLA class I expression and CD8+ lymphocyte infiltration within the tumour. With the introduction of targeted biological therapies, changing the physiological behaviour of the cancer cell may have far reaching effects on the immune response as well as outcome. Future work delineating the mechanisms underlying the local immune response could offer a novel therapeutic target for patients with a low immune cell infiltration who may have a poor outcome.

## Data Availability

The corresponding authors happy to take individual requests for data.

